# Multidimensional deprivation and brain health in older adults in rural northern Tanzania: a cross-sectional community-based study

**DOI:** 10.64898/2026.11.04.26367284

**Authors:** Sunkanmi Folorunsho, Amy Woods, Aloyce Kisoli, Grace Saria Anderson, Amour Mkubila, Malwina Niechcial, Charles Nye, Sabas M Rite, Aurelia D Malya, Sabinus L Mtenga, Hadija R Ulomi, Edward N Mbowe, Damas A Mlaki, Judith Boshe, Ssenku Safic, Hannah Humble, Connor Healy, Lachlan Fotheringham, Louise Robinson, Richard Walker, Bernard Mbwele, Raj Kalaria, Stella-Maria Paddick

**Author notes:** Department of Psychology, School of Business, Design and Social Sciences, Heriot-Watt University, Edinburgh, UK. Correspondence to: Sunkanmi Folorunsho, Department of Sociology, University of Nebraska-Lincoln, Lincoln, NE 68588, USA.

## Abstract

**Introduction:** Dementia and late-life depression are rising rapidly in sub-Saharan Africa, yet prevention frameworks remain focused on individual behaviour. Multidimensional poverty is a plausible structural determinant of brain health but is rarely assessed.

**Methods:** This cross-sectional analysis of the community-based Akili Mali study assessed adults aged 60 years and older in three rural communities in Kilimanjaro, Tanzania. Outcomes were the 15-item Geriatric Depression Scale (GDS-15), IDEA cognitive screen, CERAD ten-word list, and verbal fluency. Deprivation was measured with a nine-indicator Alkire-Foster Multidimensional Deprivation Index. Missing data were multiply imputed; models adjusted for age, sex, education, community, comorbidity, and health behaviours.

**Results:** Of 1054 participants (mean age 70.2 years [SD 9.35]; 629 [59.7%] women), 840 (81.9%) of 1026 were multidimensionally deprived. Each unit increase in deprivation score predicted a higher GDS-15 score (β 4.83, 95% CI 3.82 to 5.83), higher odds of screen-positive depression (odds ratio 17.18, 6.89 to 42.83), lower IDEA score (β −1.97, −2.97 to −0.97), and lower verbal fluency (β −2.57, −3.77 to −1.36; all p<0.001). CERAD measures and self-reported impairment showed no association. At community level this gradient inverted: the most deprived community had the lowest depression and highest cognition, yet perceived social standing did not differ.

**Conclusion:** Multidimensional deprivation was near-universal and independently associated with worse brain health. The community-level inversion, with perceived social standing equivalent across communities of unequal material circumstances, indicates that absolute deprivation alone does not explain the distribution of brain health. Relative position and community resilience are priorities for study.

**Funding:** Medical Research Council (MR/X03092X/1).

**Key messages:** *What is already known on this topic:* Multidimensional poverty has been linked to dementia in South Africa and to depressive symptoms in Kenya, but no study in East Africa has examined it against both cognition and depression in the same older population, and dementia prevention frameworks remain oriented to individually modifiable risk.

*What this study adds:* Multidimensional deprivation affected four in five older adults in rural Kilimanjaro and was independently associated with greater depressive symptom burden and poorer measured cognition, yet the relationship inverted between communities, with the most materially deprived community showing the lowest depression and the strongest cognitive performance while rating its social standing no lower than the least deprived.

*How this study might affect research, practice or policy:* Different dimensions of deprivation predict different outcomes, so anti-poverty measures intended to protect brain health may need to be dimension-specific, and the community-level inversion is a caution that deprivation data aggregated to community level can reverse the relationships operating within those communities.

## Introduction

Brain health in later life, encompassing cognitive function and mental wellbeing, is a growing global priority. Cognitive decline spans a spectrum from mild impairment to dementia,^16^ while depression is characterised by persistent low mood, loss of interest, and impaired daily functioning.^17^ Both carry substantial consequences for independence, quality of life, and mortality, share common risk factors, frequently co-occur, and may be bidirectionally related.^18,19^ The burden is shifting decisively toward the global south, and eastern sub-Saharan Africa is projected to experience one of the steepest increases in dementia cases worldwide by 2050. ^20^ In the Hai district studied here, dementia prevalence in adults aged 70 years and older rose from 6.4% in 2009–2010 to 8.9% in 2018–2019.^27^

Prevention frameworks have so far concentrated on individually modifiable risk. The 2024 Lancet Commission identified 14 modifiable risk factors accounting for approximately 45% of dementia cases worldwide.^23^ Recent work argues that this framework under-represents the realities of the global south,^21,22^ and shows that incorporating poverty, wealth shocks, income inequality, and HIV raises the potentially preventable fraction to as much as 65%.^24^ There are corresponding calls to shift prevention from individually targeted behaviour change toward structural approaches, since behaviour-based prevention places the burden on individuals and may widen disparities when better-resourced groups benefit disproportionately.^25,26^

Poverty is among the most consequential of these structural determinants, and is increasingly understood as multidimensional rather than purely economic.^1^ Individuals commonly experience overlapping social, material, and psychosocial deprivations that interact to shape health.^1,2^ This is reflected in the global Multidimensional Poverty Index adopted by the United Nations Development Programme, which assesses deprivation across health, education, and standard of living and classifies individuals as multidimensionally poor when they experience a weighted threshold of deprivations.^8,9^ Income-based measures alone capture neither the lived experience of poverty nor its consequences, whereas multidimensional measures help identify priority areas for intervention.^7^

Older adults are particularly exposed. Disadvantage accumulates across the life course, so that limited schooling, poor living conditions, and food insecurity in early life compound into later life.^3^ Older adults typically have fewer resources with which to respond to deprivation,^4^ and are more likely to live with multimorbidity and functional decline.^5,6^ Sub-Saharan Africa carries a disproportionate share of this burden, with an estimated 556 million people affected, of whom 82% live in rural areas,^10^ while experiencing some of the fastest demographic ageing globally.^13^ Evidence directly linking multidimensional deprivation to brain health in African settings remains limited. A cross-sectional study in Soweto, South Africa, found that multidimensionally poor adults aged 50 years and older had 2.31 times the risk of dementia,^50^ and work in rural Kenya has demonstrated a dose-response association between multidimensional poverty and depressive symptoms.^51^ Neither examined cognition and depression together, and to our knowledge no study in East Africa has assessed multidimensional deprivation against both domains of brain health in the same older population.

This study assessed multidimensional deprivation in older adults in the Kilimanjaro region of northern Tanzania and examined its associations with depressive symptoms and cognitive function. By identifying which dimensions of deprivation carry the greatest association with each outcome, we aimed to inform the design of targeted, multisectoral interventions.

## Methods

### Study design and setting

We conducted a cross-sectional analysis using data from the Akili Mali (Brain Wealth) study, a community-based study of older adults in the Hai district of Kilimanjaro region, northern Tanzania. Hai is a rural district on the slopes of Mount Kilimanjaro, with a population of 240 999 at the 2022 national census, 79.3% of whom live in rural areas. The majority of older residents are subsistence farmers and levels of illiteracy in this age group are high, reflecting historically limited access to schooling.^14^ Dementia prevalence in this population rose over the decade to 2019, against declines reported in several high-income settings.^27^

The study includes three rural communities stratified by geography and socioeconomic status, selected by the District Medical Office as typical exemplars of these strata: Narumu Tella, a highland community of relatively higher socioeconomic status; Orori, a neighbouring highland community of relatively lower socioeconomic status; and Sanya Station, a geographically and culturally distinct lowland community.

### Participants and recruitment

A door-to-door census was conducted in each community by balozis, elected leaders of ten households who report to the village committee, recording year of birth and sex of all residents. Individuals born in 1965 or earlier were eligible, assuming a January birth date, and all were invited through door-to-door information delivered by members of the Akili Mali Lay Advisory Board, older residents of the participating communities who advise the study and are co-authors of this paper, together with the village committee. Those too frail to attend a study centre were offered a home visit, and those too unwell to participate were excluded and referred to services.

Eligible participants were aged 60 years or older, resident in one of the three study communities, and consented to participate. Where data were implausible, field teams returned to verify and correct entries, and years of birth were revised accordingly. All residents identified by the census as born in 1965 or earlier were invited to take part, and 1054 were confirmed as aged 60 years or older on verification of year of birth. Year of birth was not known for 13 participants, whose age is therefore an estimate.

### Consent

All participants gave informed consent following a village information meeting and subsequent verbal and written information. For participants unable to read, information was read aloud by a study team member and consent was indicated using a thumbprint. Those lacking capacity to consent were included only with evidence of active willingness to participate and with consent provided by a close relative.

### Multidimensional deprivation assessment

Multidimensional deprivation was measured using a Multidimensional Deprivation Index constructed with the Alkire-Foster counting methodology.^28^ This framework is the most widely used and internationally recognised approach to multidimensional poverty measurement and underpins the global Multidimensional Poverty Index.^1,8^ The method identifies individuals experiencing overlapping deprivations across multiple domains of wellbeing.^29^

Participants were assessed against a set of nine predefined indicators, each with a deprivation cutoff, and classified as deprived or not deprived on each. Indicators were coded in binary form (1 = deprived, 0 = not deprived) and summed to give a deprivation count from 0 to 9. A normalised index score was computed as the mean of all binary indicators, producing a continuous measure between 0 and 1. Participants deprived on three or more indicators were classified as multidimensionally deprived, following the convention adopted in the global index for a nine-indicator specification.^8,28^ Those with fewer than six valid indicators were not assigned a binary classification.

The dimensions used align with the global Multidimensional Poverty Index domains of education, health, and living standards.^8^ Indicator selection was informed by the global index to facilitate comparison, while being adapted to reflect local context and available data. Within the education dimension, years of schooling and literacy capture formal attainment and current functional capability, respectively. We used an individual-level adaptation of the global index threshold of six years of schooling, and substituted literacy for school attendance to suit an older population. Within the health dimension, food insecurity and dietary diversity reflect short-term and structural aspects of food deprivation, both associated with adverse mental health and cognitive outcomes in later life.^30^ The global index uses body-mass index to indicate undernutrition, but that measure has important limitations in older populations. Living standards were assessed with five indicators. Solid cooking fuel use and inadequate housing materials follow established global index cutoffs; household air pollution from solid fuel combustion remains one of the leading environmental risk factors globally and is concentrated in low-income settings.^31^ Household crowding captures constrained living space, and absence of glass windows was used as a further marker of housing quality. Low perceived safety, although not a standard global index indicator, was included because perceived safety influences mobility, social participation, and mental health in older adults.^32^ It was defined as a score at or below the sample 25th centile, a value of 7.0 or lower on the 1 to 10 scale, because the distribution was heavily skewed toward feeling safe. The full indicator set with cutoffs, derivation and source is given in online supplemental appendix 1.

### Brain health assessment

Cognition was assessed using instruments with prior validation and normative data for Tanzania. The Identification and Intervention for Dementia in Elderly Africans (IDEA) cognitive screen total score measured general cognition. It was developed for populations with low levels of formal education and, across Nigerian and Tanzanian cohorts, showed area under the receiver operating characteristic curve above 0.9 in all settings and no evidence of educational bias.^33^ Two components were additionally analysed as full-scale scores because normative data were available: the CERAD ten-word list learning task, with minor local adaptation,^34^ and one-minute categorical verbal fluency using animal naming.

Depressive symptoms were assessed using a previously established Swahili translation of the 15-item Geriatric Depression Scale.^35^ Although the scale is designed for self-administration, high levels of illiteracy and uncorrected near vision impairment meant that for most participants it was administered by a clinical research assistant. A score of 5 or more was classified as screen-positive depression. At this cutoff, pooled sensitivity across 43 studies is 0.89 and pooled specificity 0.77.^36^ Most sociodemographic and general health measures were obtained by self-report, including history of stroke, diabetes, HIV and treated hypertension, with the exception of body-mass index and blood pressure, which were measured at assessment. Uncontrolled hypertension was defined as a measured blood pressure in the hypertensive range, averaged over three readings taken at assessment, in a participant either not taking antihypertensive medication or remaining hypertensive despite treatment. The hypertensive range was taken as a systolic pressure of 140 mmHg or higher or a diastolic pressure of 90 mmHg or higher; mean systolic pressure was 140 mmHg or higher in 718 (97.4%) of the 737 participants so classified. Perceived social standing was measured with the MacArthur ladder, on which participants placed themselves on a ten-rung ladder relative to others in their village and to others in Tanzania.

### Patient and public involvement

Older residents of the study communities were involved throughout. The Akili Mali Lay Advisory Board, whose members are drawn from the participating communities and are co-authors of this paper, contributed to the design of the study, advised on the acceptability and wording of the assessment measures, and delivered the door-to-door information that invited people to take part. Village committees and balozis supported recruitment and the community meetings at which the study was explained before consent was sought. Participants were not involved in the analysis or in drafting this manuscript. Findings will be returned to the communities through the Lay Advisory Board and the village committees.

### Statistical analysis

All analyses were done in R version 4.3.3. Continuous variables are presented as mean (SD) and categorical variables as number (%). Comparisons between deprived and non-deprived participants used Welch two-sample t tests and Pearson χZ tests. Because the three communities were purposively selected to differ in geography and socioeconomic status, characteristics and outcomes were additionally compared across sites using one-way ANOVA and χZ tests, with all three pairwise contrasts examined and p values Bonferroni-corrected.

Before finalising the index, we assessed whether any indicators carried substantially overlapping information, which would risk double-counting a single underlying deprivation. Pairwise Pearson correlations were computed across all binary indicators and variance inflation factors were estimated by regressing each indicator on all others, with a value above 5 taken as indicating problematic multicollinearity.^40^

Missingness ranged from under 1% to 13.3% across variables, being highest for verbal fluency and lowest for literacy at 0.2%, and reflecting assessment completion rather than any systematic pattern. Rather than restrict analyses to complete cases,^37^ missing data were handled by multiple imputation by chained equations, generating 20 imputed datasets with ten iterations each.^38^ Continuous variables were imputed by predictive mean matching and binary variables by logistic regression, with all exposure, outcome, and covariate variables in the imputation model. Estimates were pooled using Rubin’s rules.^39^

Associations between deprivation and each brain health outcome were estimated with the normalised index score as the exposure, using linear regression for continuous outcomes and logistic regression for binary outcomes. Model 1 was unadjusted. Model 2 adjusted for age, sex, years of education, study community with Narumu Tella as reference, multimorbidity, tobacco use, alcohol use, body-mass index, and hospitalisation in the preceding three months. Community was included as a fixed effect because the three sites were purposively selected to differ and therefore represent a structural rather than incidental source of variation. Each dimension sub-score was additionally modelled in place of the overall score, with identical adjustment.

Three prespecified sensitivity analyses were undertaken, each re-estimating every model with an alternative index: one excluding the educational deprivation indicator, compared against the standard index within an identical complete-case subsample, because years of education independently predicts cognitive test performance; one adding uncontrolled hypertension, which is used here as a marker of limited health-care access but is also an established vascular risk factor; and one weighting each dimension equally rather than each indicator equally, following the global index convention, since weighting is discretionary. Given the high prevalence of screen-positive depression, depression models were additionally re-estimated using a more conservative GDS-15 threshold of 7 or more. Statistical significance was defined as a two-sided p value below 0.05.

## Results

We included 1054 older adults from three communities: 486 (46.1%) from Narumu Tella, 353 (33.5%) from Orori, and 215 (20.4%) from Sanya Station. Mean age was 70.2 years (SD 9.35) and 629 (59.7%) were women. Educational attainment was low, with a mean of 4.09 years (SD 3.46) of formal schooling. 878 participants (85.6%) were of Chagga ethnicity and 115 (11.2%) were Maasai. Established vascular and lifestyle risk factors were common, with 254 participants (25.0%) having hypertension, 226 (25.7%) reporting ever using tobacco, and 603 (59.4%) reporting alcohol use (table 1).

**Table 1.**
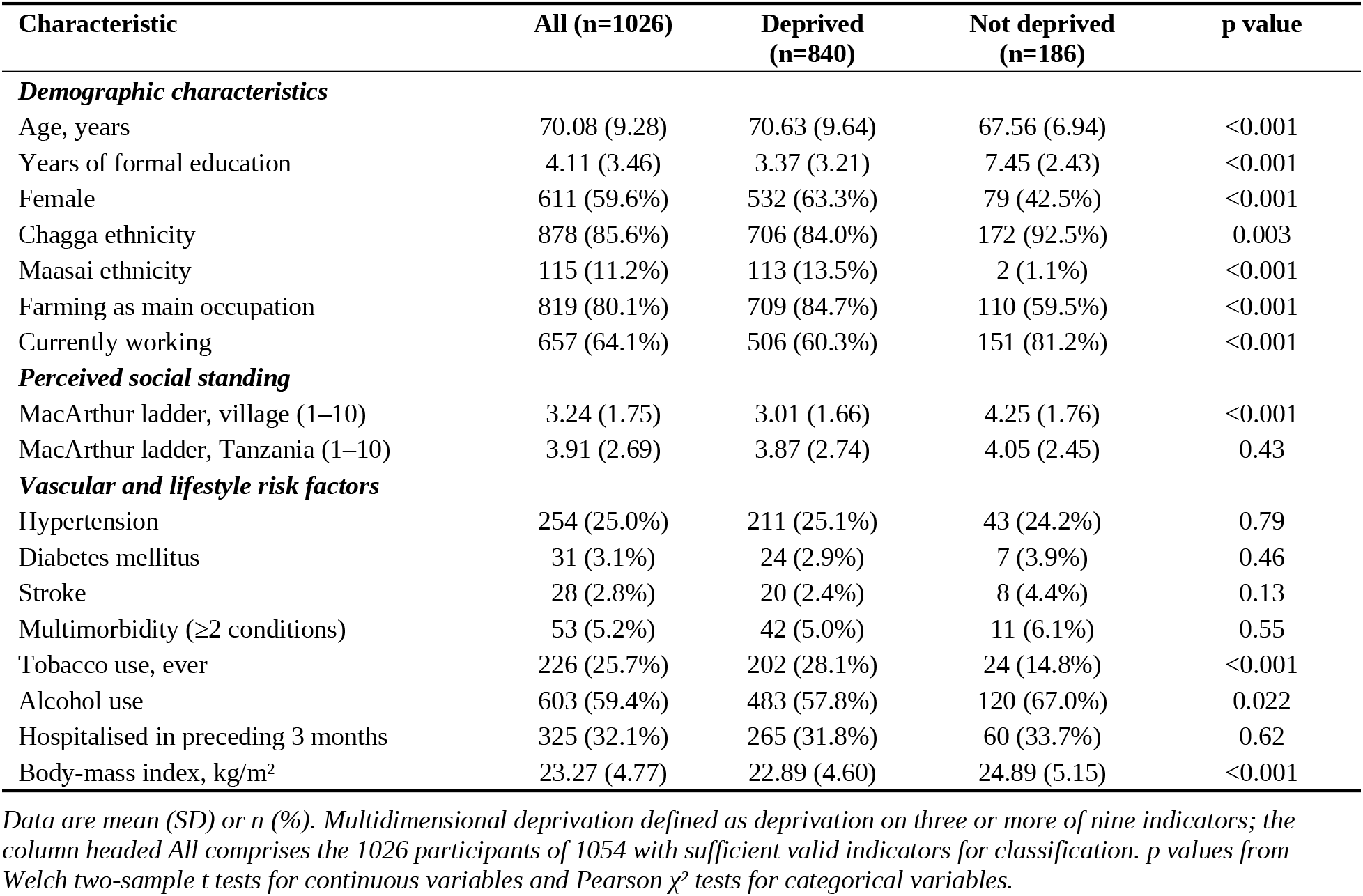
Characteristics of participants by multidimensional deprivation status.

The mean deprivation count was 4.32 of nine indicators (SD 2.00), corresponding to a mean normalised index score of 0.50 (SD 0.23). 840 (81.9%) of 1026 participants with sufficient valid indicators were classified as multidimensionally deprived. Within living standards, solid cooking fuel use was near-universal at 1000 participants (97.0%), and 557 (54.9%) lacked glass windows. Within education, 596 (56.5%) had fewer than six years of schooling and 633 (60.2%) could not read or write. Within health, 539 (53.0%) had low dietary diversity and 360 (35.0%) reported current food insecurity. Childhood food insecurity, recorded but not in the index, affected 174 (17.0%) and tracked current food insecurity (r=0.42, p<0.001). Dimension sub-scores were highest for education (0.58, SD 0.43), followed by living standards (0.48, SD 0.25) and health (0.44, SD 0.37). Sensory and vascular risk factors relevant to brain health, although not part of the core index, were common: 439 participants (46.8%) having hearing impairment, 300 (30.3%) visual impairment, and 737 (71.9%) uncontrolled hypertension (table 2).

**Table 2.** Prevalence of deprivation indicators by dimension.

| **Indicator** | **Participants assessed** | **Deprived, n** | **Deprived, %** |
|---|---|---|---|
| ***Education dimension*** |  |  |  |
| Fewer than 6 years of schooling | 1054 | 596 | 56.5 |
| Unable to read or write | 1052 | 633 | 60.2 |
| ***Health dimension*** |  |  |  |
| Current food insecurity | 1028 | 360 | 35.0 |
| Low dietary diversity (<5 food groups) | 1017 | 539 | 53.0 |
| ***Living standards dimension*** |  |  |  |
| Solid cooking fuel (wood or charcoal) | 1031 | 1000 | 97.0 |
| Inadequate housing materials | 1017 | 280 | 27.5 |
| Household crowding (>2 persons per room) | 1007 | 326 | 32.4 |
| Low perceived safety (≤25th centile) | 1022 | 267 | 26.1 |
| No glass windows | 1015 | 557 | 54.9 |
| ***Additional brain health risk factors, not in index*** |  |  |  |
| Hearing impairment (WHO grade ≥1) | 939 | 439 | 46.8 |
| Visual impairment (WHO distance score ≥1) | 990 | 300 | 30.3 |
| Uncontrolled hypertension | 1025 | 737 | 71.9 |

*Indicators coded 1 = deprived, 0 = not deprived. Participants assessed is the denominator for each indicator and varies because of item-level missing data. WHO=World Health Organization.*

No indicator pair showed correlation sufficient to indicate redundancy within the composite. The strongest association was between fewer than six years of schooling and inability to read or write (r=0.51), followed by inadequate housing materials and absence of glass windows (r=0.48). All variance inflation factors were well below the conventional threshold of 5, the highest being 1.41, so no indicators were removed or combined.

Screen-positive depression affected 710 participants (69.3%), with a mean GDS-15 score of 6.30 (SD 3.19). Multidimensionally deprived participants had higher depressive symptom burden than non-deprived participants (mean 6.58 vs 4.97; p<0.001) and were more likely to screen positive (606 [72.7%] vs 94 [52.5%]; p<0.001). They also performed worse on the IDEA cognitive screen (10.79 vs 12.32; p<0.001), CERAD list learning (11.29 vs 12.65; p<0.001), and verbal fluency (9.30 vs 11.74; p<0.001), with a consistent though marginal difference on CERAD delayed recall (2.92 vs 3.29; p=0.050). Self-reported cognitive impairment did not differ by deprivation status (448 [55.1%] vs 111 [60.7%]; p=0.17).

Deprivation differed significantly across the three communities (p<0.001), with Sanya Station most deprived (index score 0.59), followed by Orori (0.50) and Narumu Tella (0.45), and all pairwise contrasts remaining significant after Bonferroni correction. Brain health outcomes did not follow this gradient. Depression was highest in Orori (278 [80.8%]), then Narumu Tella (345 [72.5%]), and lowest in Sanya Station (87 [42.6%]), with all three pairwise contrasts significant. The same inversion was apparent for cognition, with Sanya Station participants outperforming both highland communities on every objective measure. Orori scored significantly worse than Narumu Tella on the IDEA screen (p=0.017) and CERAD list learning (p=0.003) while being only modestly more deprived. Perceived social standing, by contrast, did not differ across the three communities on either the village ladder (p=0.78) or the national ladder (p=0.79), despite the marked differences in measured deprivation (table 3).

**Table 3.** Sociodemographic characteristics, deprivation, brain health outcomes, and risk factors by community.

| **Variable** | **Narumu Tella (n=486)** | **Orori (n=353)** | **Sanya Station (n=215)** | **Overall p** | **NT vs OR** | **NT vs SS** | **OR vs SS** |
|---|---|---|---|---|---|---|---|
| ***Sociodemographic characteristics*** |  |  |  |  |  |  |  |
| Age, years | 69.24 (8.61) | 71.02 (10.19) | 70.79 (9.36) | 0.013 | 0.023 | 0.12 | >0.99 |
| Female | 278 (57.2%) | 212 (60.1%) | 139 (64.7%) | 0.18 | >0.99 | 0.19 | 0.82 |
| Years of education | 4.90 (3.33) | 4.01 (3.18) | 2.40 (3.58) | <0.001 | <0.001 | <0.001 | <0.001 |
| Farming as main occupation | 374 (77.3%) | 306 (86.7%) | 163 (76.5%) | 0.001 | 0.002 | >0.99 | 0.006 |
| ***Deprivation*** |  |  |  |  |  |  |  |
| MDI score (0–1) | 0.45 (0.21) | 0.50 (0.23) | 0.59 (0.24) | <0.001 | 0.020 | <0.001 | <0.001 |
| MDI count (0–9) | 3.96 (1.82) | 4.36 (2.02) | 5.09 (2.12) | <0.001 | 0.009 | <0.001 | <0.001 |
| Multidimensionally deprived | 370 (77.9%) | 284 (82.6%) | 186 (89.9%) | <0.001 | 0.30 | <0.001 | 0.057 |
| ***Depression*** |  |  |  |  |  |  |  |
| GDS-15 score | 6.55 (3.09) | 7.21 (2.94) | 4.18 (2.88) | <0.001 | 0.006 | <0.001 | <0.001 |
| Screen-positive depression | 345 (72.5%) | 278 (80.8%) | 87 (42.6%) | <0.001 | 0.018 | <0.001 | <0.001 |
| ***Cognition*** |  |  |  |  |  |  |  |
| IDEA score | 11.01 (3.03) | 10.32 (3.72) | 12.36 (2.85) | <0.001 | 0.017 | <0.001 | <0.001 |
| CERAD list learning | 11.38 (4.74) | 10.24 (5.02) | 13.72 (5.26) | <0.001 | 0.003 | <0.001 | <0.001 |
| CERAD delayed recall | 2.78 (2.25) | 2.50 (2.21) | 4.22 (2.47) | <0.001 | 0.24 | <0.001 | <0.001 |
| Verbal fluency | 9.56 (3.63) | 9.06 (3.68) | 10.94 (4.16) | <0.001 | 0.26 | <0.001 | <0.001 |
| Self-reported cognitive impairment | 279 (58.6%) | 178 (54.3%) | 110 (52.1%) | 0.22 | 0.66 | 0.34 | >0.99 |
| ***Perceived social standing*** |  |  |  |  |  |  |  |
| MacArthur ladder, village (1–10) | 3.22 (1.82) | 3.20 (1.59) | 3.31 (1.75) | 0.78 | >0.99 | >0.99 | >0.99 |
| MacArthur ladder, Tanzania (1–10) | 3.94 (2.66) | 3.79 (2.74) | 3.91 (2.65) | 0.79 | >0.99 | >0.99 | >0.99 |
| ***Sensory and vascular risk factors*** |  |  |  |  |  |  |  |
| Hearing impairment | 241 (52.7%) | 110 (38.3%) | 88 (45.1%) | <0.001 | <0.001 | 0.23 | 0.41 |
| Visual impairment | 155 (34.0%) | 94 (28.8%) | 51 (24.5%) | 0.038 | 0.38 | 0.043 | 0.82 |
| Uncontrolled hypertension | 384 (81.0%) | 258 (74.8%) | 95 (46.1%) | <0.001 | 0.10 | <0.001 | <0.001 |

*Data are mean (SD) or n (%). Overall p values from one-way ANOVA for continuous variables and Pearson χ² tests for categorical variables. Pairwise p values are Bonferroni-corrected for three comparisons. MDI=Multidimensional Deprivation Index. GDS-15=15-item Geriatric Depression Scale. IDEA=Identification and Intervention for Dementia in Elderly Africans. NT=Narumu Tella. OR=Orori. SS=Sanya Station.*

### Multidimensional deprivation and brain health in rural northern Tanzania

In adjusted models, each unit increase in normalised deprivation score, equivalent to moving from deprivation on no indicators to deprivation on all nine, was associated with a 4.83 point increase in GDS-15 score (95% CI 3.82 to 5.83; p<0.001) and with 17.18 times higher odds of screen-positive depression (95% CI 6.89 to 42.83; p<0.001). Expressed per additional deprivation, this corresponds to an odds ratio of 1.37 (95% CI 1.24 to 1.52). The adjusted estimates exceeded the unadjusted estimates for both depression outcomes. Associations with cognition remained significant after adjustment for global cognitive screening (β −1.97, 95% CI −2.97 to −0.97; p<0.001) and verbal fluency (β −2.57, 95% CI −3.77 to −1.36; p<0.001), but not for CERAD list learning (β −0.36; p=0.66) or delayed recall (β −0.16; p=0.67). Self-reported cognitive impairment showed no association in either model (table 4).

**Table 4.** Associations between multidimensional deprivation and brain health outcomes.

| **Outcome** | **Model 1 (95% CI)** | **p value** | **Model 2 (95% CI)** | **p value** |
|---|---|---|---|---|
| ***Depression*** |  |  |  |  |
| GDS-15 score, β | 3.21 (2.36 to 4.07) | <0.001 | 4.83 (3.82 to 5.83) | <0.001 |
| Screen-positive depression, OR | 5.49 (2.96 to 10.19) | <0.001 | 17.18 (6.89 to 42.83) | <0.001 |
| ***Cognition*** |  |  |  |  |
| IDEA score, β | −4.12 (−5.03 to −3.21) | <0.001 | −1.97 (−2.97 to −0.97) | <0.001 |
| CERAD list learning, β | −2.33 (−3.68 to −0.98) | <0.001 | −0.36 (−1.98 to 1.25) | 0.66 |
| CERAD delayed recall, β | −0.83 (−1.46 to −0.20) | 0.010 | −0.16 (−0.92 to 0.59) | 0.67 |
| Verbal fluency, β | −5.69 (−6.78 to −4.61) | <0.001 | −2.57 (−3.77 to −1.36) | <0.001 |
| Self-reported impairment, OR | 0.65 (0.37 to 1.14) | 0.13 | 1.02 (0.48 to 2.17) | 0.96 |

*Estimates are per one unit increase in normalised deprivation score (range 0 to 1). Model 1 is unadjusted. Model 2 is adjusted for age, sex, years of education, community, multimorbidity, tobacco use, alcohol use, body-mass index, and hospitalisation in the preceding three months. β=unstandardised regression coefficient from linear regression. OR=odds ratio from logistic regression. Estimates pooled across 20 imputed datasets using Rubin's rules. n=1054.*

Dimension-specific models showed contrasting patterns for the two outcomes. For depressive symptoms, living standards deprivation was the strongest predictor (β 2.75; p<0.001), ahead of health (β 1.77; p<0.001) and education (β 1.01; p=0.011). For global cognition this ordering was reversed, with education the dominant dimension (β −1.27; p=0.001) and health (β −0.68; p=0.007) and living standards (β −0.82; p=0.029) showing smaller but still significant associations (table 5).

**Table 5.**
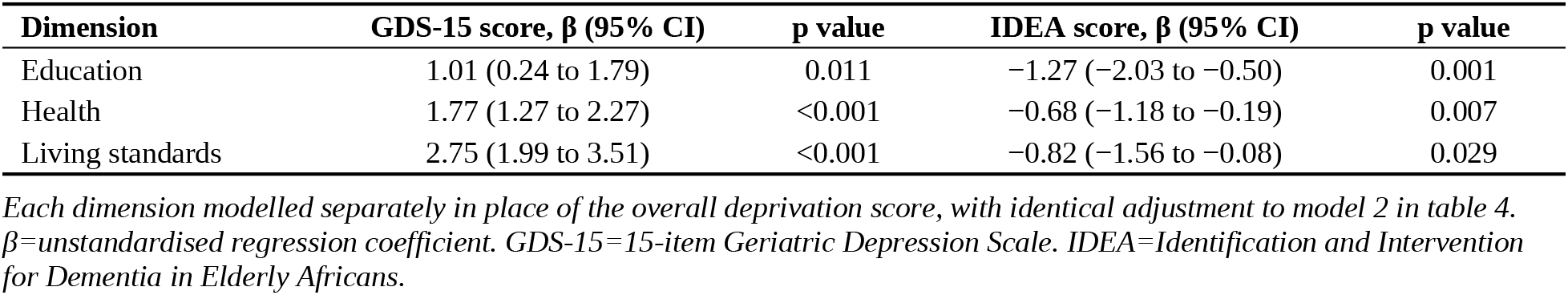
Associations between deprivation dimensions and brain health outcomes.

Excluding the education indicator from the index, adding uncontrolled hypertension to it, and re-estimating every model with equal-dimension rather than equal-indicator weighting each left the associations unchanged in direction, magnitude, and significance (online supplemental table S1).

## Discussion

In this community-based study of 1054 older adults in rural northern Tanzania, multidimensional deprivation was the norm rather than the exception, with more than four in five participants meeting the threshold for multidimensional poverty. Higher index scores predicted greater depressive symptom burden, markedly increased odds of screen-positive depression, and poorer performance on global cognitive screening and verbal fluency, after adjustment for age, sex, education, community, comorbidity, and health behaviours. Three findings merit particular attention: the depression association strengthened rather than attenuated on adjustment; deprivation tracked measured cognition but bore no relationship to participants’ own reports of difficulty; and the most deprived community had the best brain health outcomes, inverting the individual gradient.

The magnitude of the depression association is substantial, corresponding to an odds ratio of 1.37 for each additional deprivation and to a 17.2-fold difference in odds across the full range of the index after full adjustment. That the association strengthened on adjustment requires explanation rather than dismissal. Sanya Station combined the highest material deprivation with by far the lowest depression prevalence, so in unadjusted analysis this community-level pattern operated against the individual-level association and suppressed it; only when community entered the model did the underlying relationship emerge fully. This is substantive rather than technical, because it shows that aggregate comparisons across communities can obscure, and even reverse, relationships operating within them, a caution directly relevant to how population-level deprivation data are read in low-resource settings. That living standards deprivation was the strongest predictor of depressive symptoms, ahead of health and education, suggests that the immediate material conditions of daily life may bear more directly on mood in later life than educational history.

Our observed prevalence of screen-positive depression, at 69.3%, is high and warrants careful framing. Prevalence estimates for late-life depression in sub-Saharan Africa vary by an order of magnitude according to ascertainment method, with the Geriatric Depression Scale consistently yielding higher pooled prevalence than structured diagnostic interviews.^15,41^ At the cutoff used here, pooled specificity of 0.77 implies that a meaningful proportion of those screening positive would not meet diagnostic criteria.^36^ Regional comparison is instructive: in neighbouring Moshi district a community study using the same instrument reported depressive symptoms in 44.4%,^52^ whereas in Hai district structured DSM-IV assessment yielded 21.2%.^53^ Our figure should therefore be read as symptom burden captured by a screening instrument rather than the prevalence of major depressive disorder. Applying a more conservative threshold of 7 or more reduced prevalence to 46.7% and strengthened rather than weakened the association with deprivation (odds ratio 18.52, 95% CI 8.24 to 41.66; p<0.001), indicating that the finding is not an artefact of a permissive cutoff.

Associations with global cognitive screening and verbal fluency were robust to full adjustment, whereas those with CERAD list learning and delayed recall were not. Verbal fluency draws on lexical access, executive control, and processing speed, and may be a more sensitive marker of early or diffuse cognitive change than verbal learning and recall, which can show floor effects in populations with limited formal schooling. That the memory associations attenuated specifically after adjustment for age, education, and community suggests these variables, rather than deprivation itself, account for the crude differences initially observed. Education emerged as the dominant deprivation dimension for cognition, consistent with cognitive reserve theory.^42^ This raises a methodological concern, since educational deprivation contributes to the index while years of education independently predicts cognitive test performance. Removing the education indicator from the composite left the associations with global cognition and verbal fluency essentially unchanged, indicating that the relationship is not reducible to educational attainment, and the IDEA screen itself showed no evidence of educational bias across Nigerian and Tanzanian cohorts.^33^ The absence of any association with self-reported cognitive impairment, set against consistent associations with objective performance, carries practical weight, because it indicates that subjective cognitive complaint is a poor proxy for measured cognition in this setting and that screening approaches relying on self-report in low-resource contexts may systematically fail to identify those most affected.

The most theoretically challenging finding is the inversion between material deprivation and brain health at community level. Sanya Station was the most deprived community overall and on most individual indicators, yet had the lowest depression prevalence and the strongest cognitive performance, while Orori, intermediate in overall material deprivation, had the worst outcomes on most measures and performed significantly worse than its immediate highland neighbour Narumu Tella on both depression and cognition. This pattern resists explanation through absolute material deprivation alone but is consistent with relative deprivation theory, which holds that health is shaped not only by absolute resources but by an individual’s standing relative to a salient reference group, with adverse comparison generating psychosocial stress.^43^ Two pathways are conventionally proposed, one material and one psychosocial, the latter operating through chronic stress and cumulative allostatic load.^44^ Our measure of perceived social standing bears directly on this reading: within the the village (Spearman r −0.35, p<0.001) and, more weakly, nationally (r −0.08, p=0.029), so the ladder does track measured deprivation at the individual level. Across communities, however, perceived standing was statistically indistinguishable: village ladder means were 3.22 in Narumu Tella, 3.20 in Orori, and 3.31 in Sanya Station (p=0.78), and national ladder means were 3.94, 3.79, and 3.91 respectively (p=0.79), even though measured deprivation differed significantly between every pair of communities. That older adults in the most materially deprived community rate their social position no lower than those in the least deprived is itself consistent with a relative rather than absolute standard of comparison: if people judge their position chiefly against neighbours rather than against the country, perceived standing will be approximately equalised even where objective circumstances diverge. The salient comparison for Orori residents is Narumu Tella, an adjacent community selected precisely because it is better off, whereas Sanya Station is geographically and culturally distinct from both highland communities, so its residents’ reference group is likely internal to a community in which deprivation is widely shared. Sanya Station also differs in topography, natural resources and ethnic composition, being the only community with a substantial Maasai population, and these differences may bear on both outlook and assessment. Language is the most immediate concern, since all instruments were administered in Swahili, but it does not appear to explain the pattern: confidence speaking Swahili did not differ across the three communities (96.3%, 96.3% and 93.0% respectively; p=0.11), and Sanya Station residents were the most likely of the three to give Swahili as their preferred language (87.4% versus 79.2% and 49.0%; p<0.001), so any language disadvantage would work against the better performance observed there. We emphasise that the ladder captures self-rated position rather than explicit comparison with a specified reference group, so this interpretation remains hypothesis-generating. It nonetheless converges with evidence that structural inequality shapes brain outcomes beyond individual characteristics, including multi-country work in which income inequality predicted brain dynamics more strongly than individual demographic or cognitive factors.^45^ An alternative and complementary possibility is that factors operating at individual, social and community level in Sanya Station attenuate the effect of material deprivation. Social cohesion, shared cultural identity, intergenerational support, and dietary or activity differences between lowland and highland settings are plausible candidates, but were not measured here.^46,47^ These findings speak to a growing argument that dementia and brain health prevention frameworks must extend beyond individual behaviour change to address structural determinants.^25,26^ Our data are consistent with the argument that the 2024 Lancet Commission framework under-represents the realities of the global south,^23,24^ and extend the small African evidence base, which has linked multidimensional poverty to dementia in South Africa and to depressive symptoms in Kenya, by examining cognition and depression together in the same East African population, a region projected to experience one of the steepest global increases in dementia cases by 2050.^20^ That living standards deprivation predicted depression most strongly while educational deprivation predicted cognition most strongly suggests that different dimensions of poverty may call for different responses, and the balance is likely to differ by setting: individual behaviour change may carry most of the preventable fraction where living conditions and health care are already secure, whereas in lower-income settings structural measures such as access to care, housing and food security plausibly come first, with behaviour change adding to rather than substituting for them. We also note that 737 participants (71.9%) had uncontrolled hypertension, at once a marker of limited health-care access and an established vascular risk factor for cognitive decline, and one for which prevention pathways are well described.^23,48,54^ Our sensitivity analysis confirmed that this dual role does not drive our findings, but the figure itself is a substantial and directly addressable gap.

This study has several strengths, including a large community-based sample with high participation, assessment instruments validated for the local setting, a theoretically grounded deprivation measure, principled handling of missing data, and prespecified sensitivity analyses addressing the main structural concerns with the composite. Several limitations must be acknowledged. The cross-sectional design precludes inference about temporal ordering or causal direction, and depression and cognitive impairment may plausibly contribute to deprivation as well as result from it.^18^ Deprivation was measured at a single time point, so we cannot distinguish chronic from recently acquired disadvantage, a meaningful limitation given that cumulative life-course disadvantage is central to the conceptual model motivating this work. ^3^ Both the GDS-15 and the IDEA are screening instruments rather than diagnostic assessments. Perceived social standing was measured only as self-rated position on the MacArthur ladder, which does not identify the reference group against which participants compare themselves, so relative deprivation could not be tested directly. Several index indicators, notably food insecurity and perceived safety, are self-reported as is the depression measure, raising the possibility of common method bias;^49^ the convergence of the depression finding with objectively measured cognitive outcomes offers partial reassurance but cannot exclude this. Several standard global index indicators, including sanitation, drinking water, asset ownership, and child mortality, could not be included because the corresponding data were unavailable, so our index is an adapted rather than complete implementation. Residential history would help in interpreting the community-level contrast, but the item was administered to only a small subset of participants and almost entirely in one community, so it could not be compared across sites. Year of birth was not known for a small number of participants, whose age was estimated, and a small number of recorded ages at the extreme upper end of the distribution should be treated as approximate. One finding remains unexplained: net of deprivation and other covariates, more years of education was associated with modestly higher depressive symptom scores, which runs counter to the typically protective association reported in the literature. This may reflect heightened awareness of relative disadvantage, differential item interpretation, or residual confounding, and merits dedicated investigation. Finally, the three communities lie within a single district of one Tanzanian region and were purposively selected as exemplars of highland and lowland, higher and lower socioeconomic status settings, so the community-level inversion we observed is specific to this configuration and should not be assumed to be generalisable without replication.

## Conclusion

Multidimensional deprivation was highly prevalent among older adults in rural northern Tanzania and was independently associated with greater depressive symptom burden and poorer objectively measured cognition. Different dimensions of deprivation predicted different outcomes, which suggests that anti-poverty interventions targeting brain health may need to be dimension-specific rather than undifferentiated. The inversion of this relationship at the community level, where the most materially deprived community showed the best outcomes while rating its social position no lower than the least deprived, indicates that absolute deprivation alone cannot explain the distribution of brain health and points toward relative position within a salient reference group, and toward unmeasured resilience factors in the least materially advantaged community, as candidate explanations meriting dedicated study. These findings support calls to broaden dementia and brain health prevention frameworks beyond individual behaviour change to encompass the structural conditions in which older adults live.

## Supporting information

Supplemental table S1: sensitivity analyses using alternative index specifications

## Acknowledgements

We thank the Akili Mali Lay Advisory Board, the village committees and balozis of the participating communities, and all study participants.

## Contributors

SMP conceived the Akili Mali study and leads it. SF, AW and SMP designed this analysis. SF prepared the analytical dataset, did the statistical analysis and drafted the manuscript, supervised by SMP. GSA, AKi, AMk, JB, HH, CH, LF, CN, MNM, BM and DAM contributed to data collection and to the conduct of the study. SMR, ADM, SLM, HRU and ENM, as members of the Akili Mali Lay Advisory Board, contributed to the design of the study, advised on the acceptability and wording of the assessment measures and supported recruitment. RNK, RW, LR and SS contributed to the interpretation of the findings. All authors revised the manuscript critically for important intellectual content and approved the final version. SF is guarantor.

## Patient consent for publication

Not applicable.

## Ethics approval

This study involves human participants and was approved by Newcastle University, the Mbeya College of Health Sciences Institutional Review Board, and the Tanzanian National Research Ethics Committee. Participants gave informed consent to participate in the study before taking part.

## Data availability statement

Data are available on reasonable request. The analytical dataset and the code that generates every result reported here are available from the corresponding author on reasonable request, subject to the approval of the Akili Mali study steering group.

## Supplemental material

Online supplemental table S1: sensitivity analyses using alternative index specifications. Online supplemental appendix 1: deprivation indicators, cutoffs and derivation.

