## Supplemental table S1: sensitivity analyses using alternative index specifications for "Multidimensional deprivation and brain health in older adults in rural northern Tanzania: a cross-sectional community-based study"

| Outcome | Standard index estimate (95% CI) | Alternative index estimate (95% CI) | p value |
|---|---|---|---|
| ***Panel A: index excluding the education indicator (complete-case analysis)*** | | | |
| IDEA score, β | −1.99 (−3.09 to −0.90) | −1.74 (−2.72 to −0.75) | <0.001 |
| CERAD list learning, β | −0.63 (−2.48 to 1.21) | −0.32 (−1.98 to 1.34) | 0.71 |
| CERAD delayed recall, β | −0.16 (−1.06 to 0.74) | −0.02 (−0.83 to 0.78) | 0.95 |
| Verbal fluency, β | −2.13 (−3.54 to −0.72) | −1.85 (−3.12 to −0.57) | 0.005 |
| ***Panel B: index including uncontrolled hypertension (multiply imputed)*** | | | |
| GDS-15 score, β | 4.83 (3.82 to 5.83) | 4.93 (3.83 to 6.04) | <0.001 |
| Screen-positive depression, OR | 17.18 (6.89 to 42.83) | 18.76 (6.95 to 50.66) | <0.001 |
| IDEA score, β | −1.97 (−2.97 to −0.97) | −1.88 (−2.97 to −0.79) | <0.001 |
| CERAD list learning, β | −0.36 (−1.98 to 1.25) | 0.01 (−1.76 to 1.77) | >0.99 |
| CERAD delayed recall, β | −0.16 (−0.92 to 0.59) | 0.10 (−0.72 to 0.93) | 0.81 |
| Verbal fluency, β | −2.57 (−3.77 to −1.36) | −2.37 (−3.69 to −1.06) | <0.001 |
| Self-reported impairment, OR | 1.02 (0.48 to 2.17) | 0.96 (0.43 to 2.15) | 0.92 |
| ***Panel C: dimension-weighted index (multiply imputed)*** | | | |
| GDS-15 score, β | 4.83 (3.82 to 5.83) | 4.87 (3.85 to 5.89) | <0.001 |
| Screen-positive depression, OR | 17.18 (6.89 to 42.83) | 19.22 (7.64 to 48.33) | <0.001 |
| IDEA score, β | −1.97 (−2.97 to −0.97) | −2.25 (−3.27 to −1.23) | <0.001 |
| CERAD list learning, β | −0.36 (−1.98 to 1.25) | −0.60 (−2.25 to 1.04) | 0.47 |
| CERAD delayed recall, β | −0.16 (−0.92 to 0.59) | −0.24 (−1.01 to 0.53) | 0.54 |
| Verbal fluency, β | −2.57 (−3.77 to −1.36) | −2.36 (−3.57 to −1.14) | <0.001 |
| Self-reported impairment, OR | 1.02 (0.48 to 2.17) | 1.11 (0.52 to 2.39) | 0.78 |

*Panel A compares the standard nine-indicator index against an eight-indicator index excluding educational deprivation, within identical complete-case subsamples (n=741 for IDEA score, 751 for CERAD list learning, 751 for CERAD delayed recall, and 690 for verbal fluency); p values shown are for the alternative index. Panel B compares the standard nine-indicator index against a ten-indicator index additionally including uncontrolled hypertension, using multiple imputation (n=1054); p values shown are for the alternative index. Panel C compares the standard nine-indicator index, in which every indicator is weighted equally, against an index in which each of the three dimensions is weighted equally and its weight divided among its indicators, using multiple imputation (n=1054); the two versions of the index correlate at r=0.96 and p values shown are for the alternative index. All models adjusted as for model 2 in table 4. β=unstandardised regression coefficient. OR=odds ratio.*
